## Supplemental Figures for "Estimating the Effects of Legalizing Recreational Cannabis on Newly Incident Cannabis Use"

**Supporting Information**

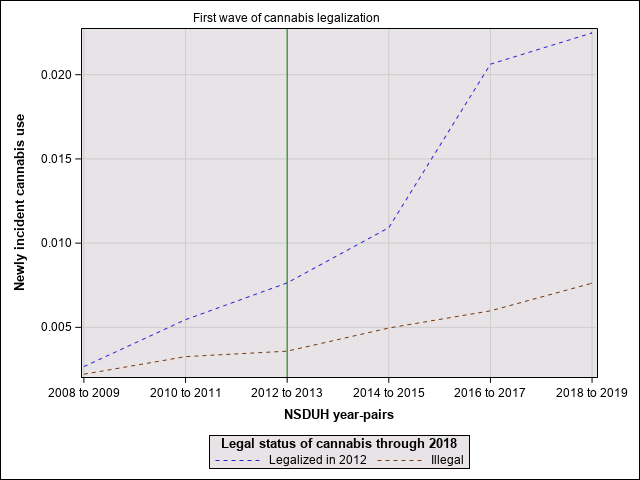

Fig S1. Cannabis incidence in 21 and older age group, first wave legalizing states vs untreated states.

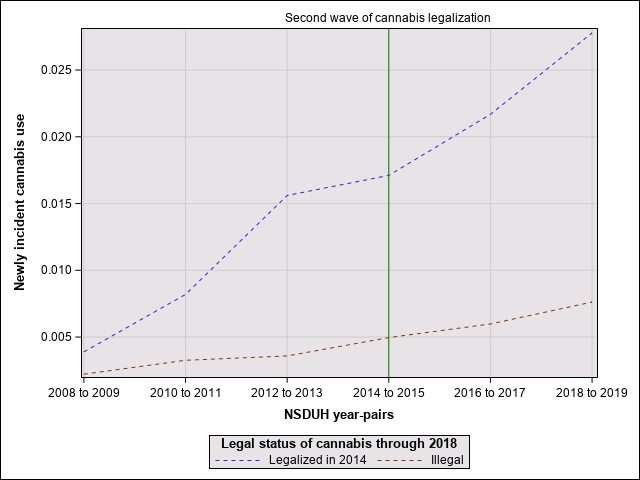

Fig S2. Cannabis incidence in 21 and older age group, second wave legalizing states vs untreated states.

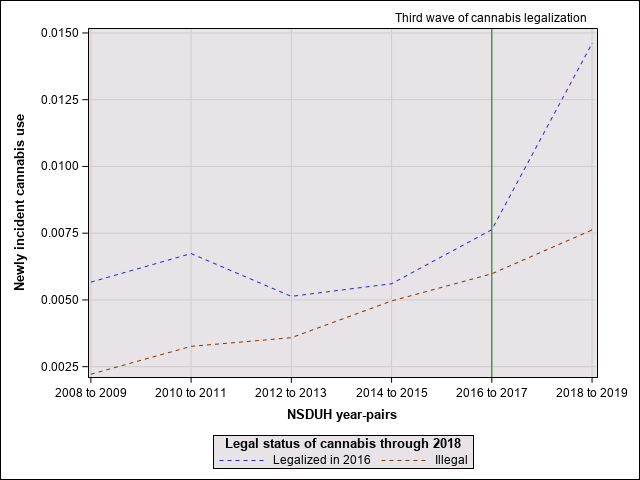

Fig S3. Cannabis incidence in 21 and older age group, third wave legalizing states vs untreated states.

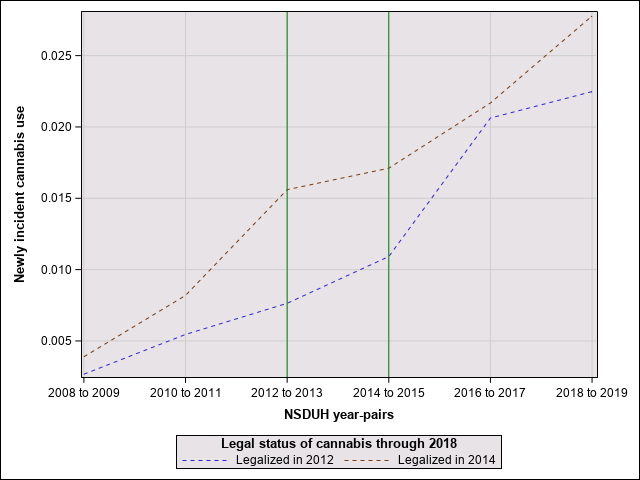

Fig S4. Cannabis incidence in 21 and older age group, first wave legalizing states vs third wave legalizing states.

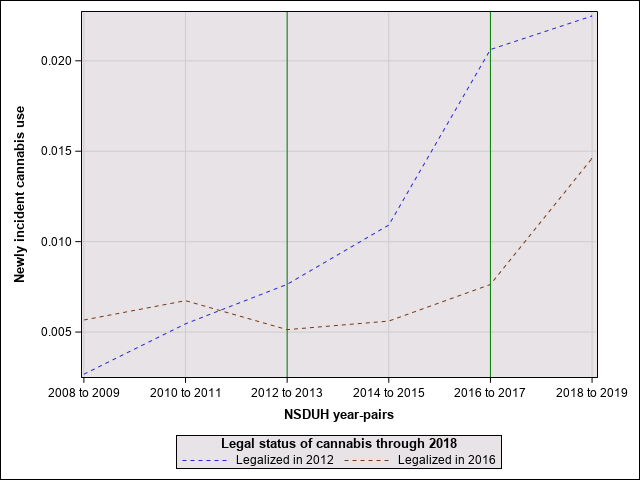

Fig S5. Cannabis incidence in 21 and older age group, second wave legalizing states vs third wave legalizing states.

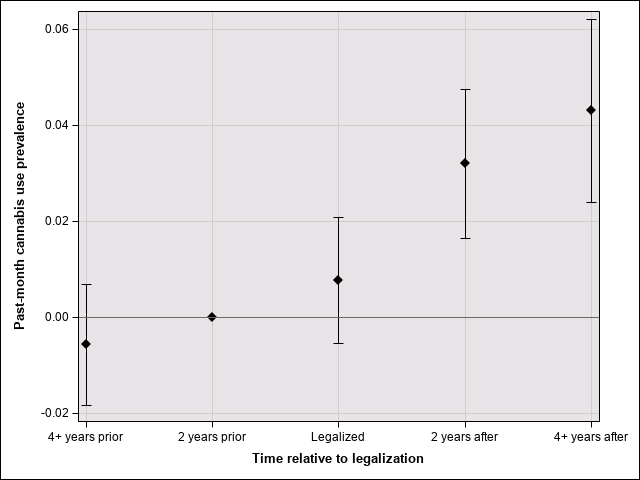

Fig S6. Effect of time since cannabis legalization on past month cannabis prevalence in the 21 and older age group.

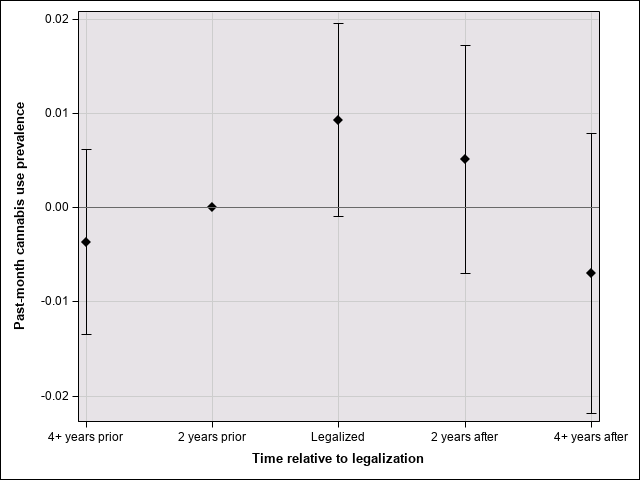

Fig S7. Effect of time since legalization on past-month cannabis prevalence in the 12-to-20-age-group.

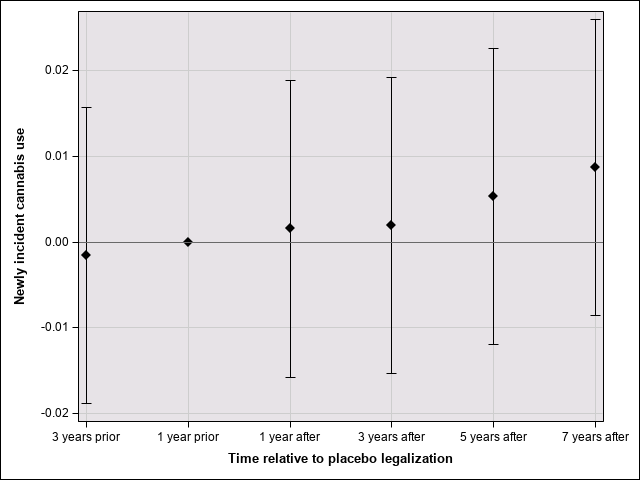

Fig S8. Placebo effect of time since cannabis legalization on cannabis incidence in the 21 and older age group.

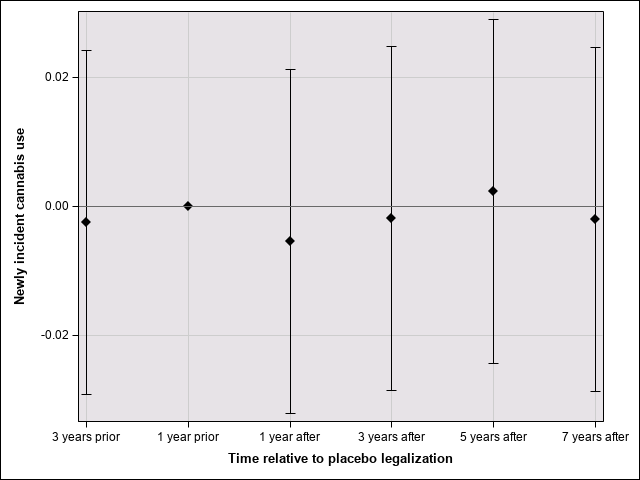

Fig S9. Placebo effect of time since cannabis legalization on cannabis incidence in the 12-to-20-age-group.
